# Trials that turn from retrospectively registered to prospectively registered: A cohort study of ‘retroactively prospective’ clinical trial registration using history data

**DOI:** 10.1101/2022.10.25.22281505

**Authors:** Martin R. Holst, Benjamin G. Carlisle

## Abstract

**Background:** Prospective clinical trial registration is a methodological best practice, a moral requirement under the Declaration of Helsinki, and in many cases legally required. The US clinical trials register ClinicalTrials.gov allows for changes to a clinical trial registry entry at any time, including changes to the clinical trial start date, making it possible for a trial that was registered after the enrolment of the first patient (retrospective registration) to retroactively change its start date to a later one, making it appear to be prospectively registered.

**Methods:** Using the novel R package cthist, we downloaded clinical trial history data for all clinical trials with a first registration date in the year 2015.

**Findings:** We found 235 clinical trials to be ‘retroactively prospective’, comprising 2.0% of all clinical trials first registered on ClinicalTrials.gov in 2015 and 3.9% of all prospectively registered trials. Among the 113 retroactively prospective clinical trials with an accompanying publication, 10.6% explicitly stated in the publication that they had been prospectively registered.

**Interpretation:** While the rate of retroactively prospective trial registration is lower than hypothesized, the existence of this issue has implications for the review and conduct of clinical trials, and it can be detected using fully automated and freely available tools. This is the first analysis of the potentially questionable research practice that we call ‘retroactively prospective’ trial registration.

**Funding:** This work was funded by internal funding at QUEST Center for Responsible Research.

## Introduction

Clinical trials provide an important foundation of the medical evidence base. Prospective registration of clinical trials – that is, registration in a public database like ClinicalTrials.gov before the first patient is enrolled – has become mandated by different regulations in order to facilitate a complete and unbiased publication record, restrict researcher degrees of freedom, inform the public, and fulfill ethical obligations (1–4). In 2005, the International Committee of Medical Journal Editors (ICMJE) implemented a requirement that all clinical trials be registered before the first patient is enrolled in order to publish in an ICMJE journal (5). The US Food and Drug Administration Amendments Act of 2007 Sec. 801 requires that certain clinical trials be registered not later than 21 days after the first patient is enrolled (6). The CONSORT Explanation and Elaboration documentation in 2010 also mentions prospective registration of a clinical trial before the first patient is enrolled (7). Finally, the Declaration of Helsinki states that ‘[e]very research study involving human subjects must be registered in a publicly accessible database before recruitment of the first subject’ (8).

Whether a trial is registered prospectively can be determined by looking at the start date (i.e., the date the first patient is enrolled in the study) of the trial compared to the registration date (i.e., the date the trial registration is submitted to the registry): If the start date is equal to or is after the registration date, then the trial has been registered prospectively. If the start date is before the registration date, then the trial has been registered retrospectively. It is important to note, however, that ClinicalTrials.gov allows investigators to update the start date of their trial at any time. This does not remove the original trial registry entry version, which can be found by visiting the historical changes page for the trial in question, and which is in principle accessible to anyone. This information could so far not be retrieved through most tools provided for downloading ClinicalTrials.gov records for larger-scale analysis, as the data did not include historical versions. This has changed with the release of the cthist R package (9), which was developed for this project.

In this study, we aimed to investigate and describe changes to historical versions of clinical trial registry entries for a one-year cohort of ClinicalTrials.gov records. Our main aim is to determine the frequency with which recorded start dates are changed in a manner that would obscure the fact that the trials were originally registered retrospectively. We defined this to be ClinicalTrials.gov records whose first reported start date (post-launch) indicates retrospective registration (i.e., the start date is before the registration date), but where the start date was pushed forwards such that at 5 years after first registration, the trial registry entry indicates prospective registration (i.e., the start of that trial is equal to or dated after the registration date, see Figure 1). Thus, our primary objective was to measure the number and proportion of these so-called ‘retroactively prospective’ trial registrations.

**Figure 1.**
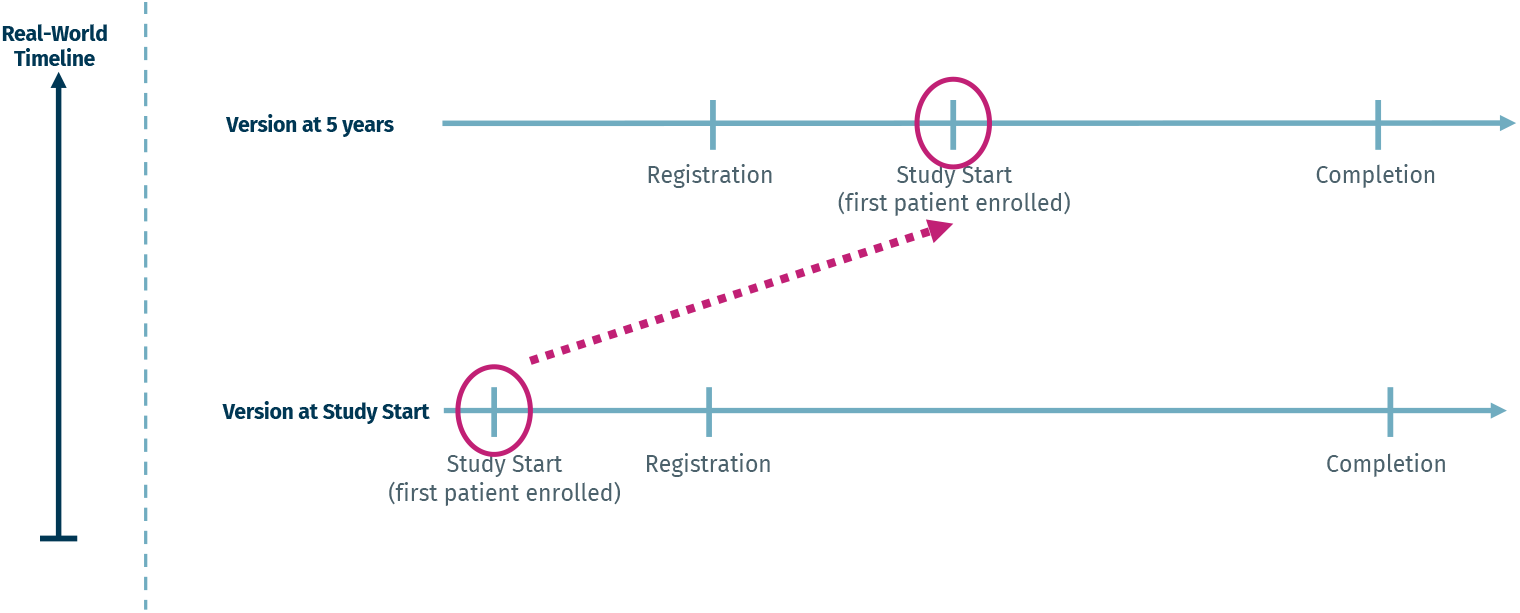
Example for ‘retroactively prospective’ trial registration. By the time the first patient is enrolled, the trial registry record indicates retrospective registration, but by 5 years, the start date has been pushed forward, indicating prospective registration.

As secondary objectives, we aimed to measure further frequencies and proportions, for example the number of trials that stay either retrospective or prospective from the beginning. We also describe whether these dynamics change by phase number or medical field. Furthermore, we estimate the rate of concordance between the first post-launch start date of clinical trials and the start date that is reported in the respective journal publication. Additionally, we aim to describe how many trials report a change to their start dates, as well as the timepoint of the changes.

## Methods

Details of our methods can be found in our preregistered trial protocol, which is available on the Open Science Framework (OSF, https://osf.io/82vfn/). The most recent version of the protocol also lists all deviations from the original study plan, including the rationale behind the changes. Our manuscript adheres to the STROBE Checklist for cross-sectional studies (10).

### Data sources and sampling

We downloaded the national clinical trial (NCT) numbers for all interventional clinical trial records with a registration date between 1 January 2015 and 31 December 2015 from the ClinicalTrials.gov website. For this set of trials, we extracted the complete historical versions using the newly developed cthist R package (9), a tool designed to automatically retrieve historical versions of clinical trial registry records. Using the downloaded historical versions data, we excluded trials that were not Terminated or Completed by 5 years after their first registration date. To ensure equal follow-up, we also excluded all historical versions that were posted more than five years after registration.

### Automated analyses

We determined whether trials examined one of four broad medical fields (cancer, cardiovascular, neurological, pain) by searching for the respective indications on ClinicalTrials.gov and integrating the results into our database. We chose these broad fields because they cover a large area of medical research, are broad enough to provide a good sample from each, and have distinct goals and methods. We performed automated analyses to determine the registration status (i.e., retrospective or prospective) of the trials at study start (first registered version after the trial is set to Recruiting, Enrolling by invitation, Active, not recruiting, Completed, or Terminated) and at 5 years (latest historical version within the 5-year range). Using these data, we were able to directly identify trials that constitute ‘retroactively prospective’ trial registration. Due to changes in policy on ClinicalTrials.gov, some of the study start dates were rounded to the month in the registry entry, while others were exact. We accounted for this by performing separate calculations: If the precision of the start date was only to the month, we used rounded dates for both the start date and the registration date; if the precision of the start date was to the day, we based our calculation on precise dates.

### Matching trials to publications

All published ‘retroactively prospective’ trials and a random sample of 200 published trials that were originally prospectively or retrospectively registered, and remained so until 5 years of follow-up, were selected for human ratings. Using the PubMed application programming interface (API), these trials were automatically matched with journal publications indexed in PubMed that listed the corresponding ClinicalTrials.gov NCT number in their ‘secondary identifier’ field or in the text of the title/abstract.

### Manual assessments

The downloaded historical versions data were combined with manually extracted data using Numbat Systematic Review Manager (11). Each corresponding publication was assessed by two of three independent extractors (MRH, BGC, SY). We first checked whether the publication was indeed a results publication for the respective trial. Then, we used the full text to extract start dates, end dates, and whether the trial mentioned ‘prospective’ or ‘retrospective’ registration. We also extracted whether any change to the start date was mentioned in the publication. Any discrepancies were discussed and resolved.

### Sample size rationale and calculation

Because most of our analyses can be calculated using data that do not require human curation, we saturated our sampling frame and took all eligible trials that meet our inclusion criteria. For our manually assessed data, we calculated that a sample of at least 200 observations per group was expected to provide 80% power to detect a difference in proportions of 10% and 20% with a significance level of 0·05.

### Data and code availability

Our code is available via an online repository (https://codeberg.org/bgcarlisle/RetroProspectiveCTR). Our dataset, which includes both the publicly available and the manually extracted data, is available in our OSF repository.

### Hypotheses

(1) We hypothesized that there would be a rate of ‘retroactively prospective’ registrations greater than 5%. (2) We hypothesized that among clinical trials with ‘retroactively prospective’ registration, there would be a lower concordance between the originally registered study start date (post-launch) and the published start date, compared to control trials.

## Results

Our flow of included trials is shown in Figure 2. We first downloaded the historical versions data of all clinical trials registered on ClinicalTrials.gov in 2015 on 4 April 2022 (*147,377 historical versions*).

**Figure 2.**
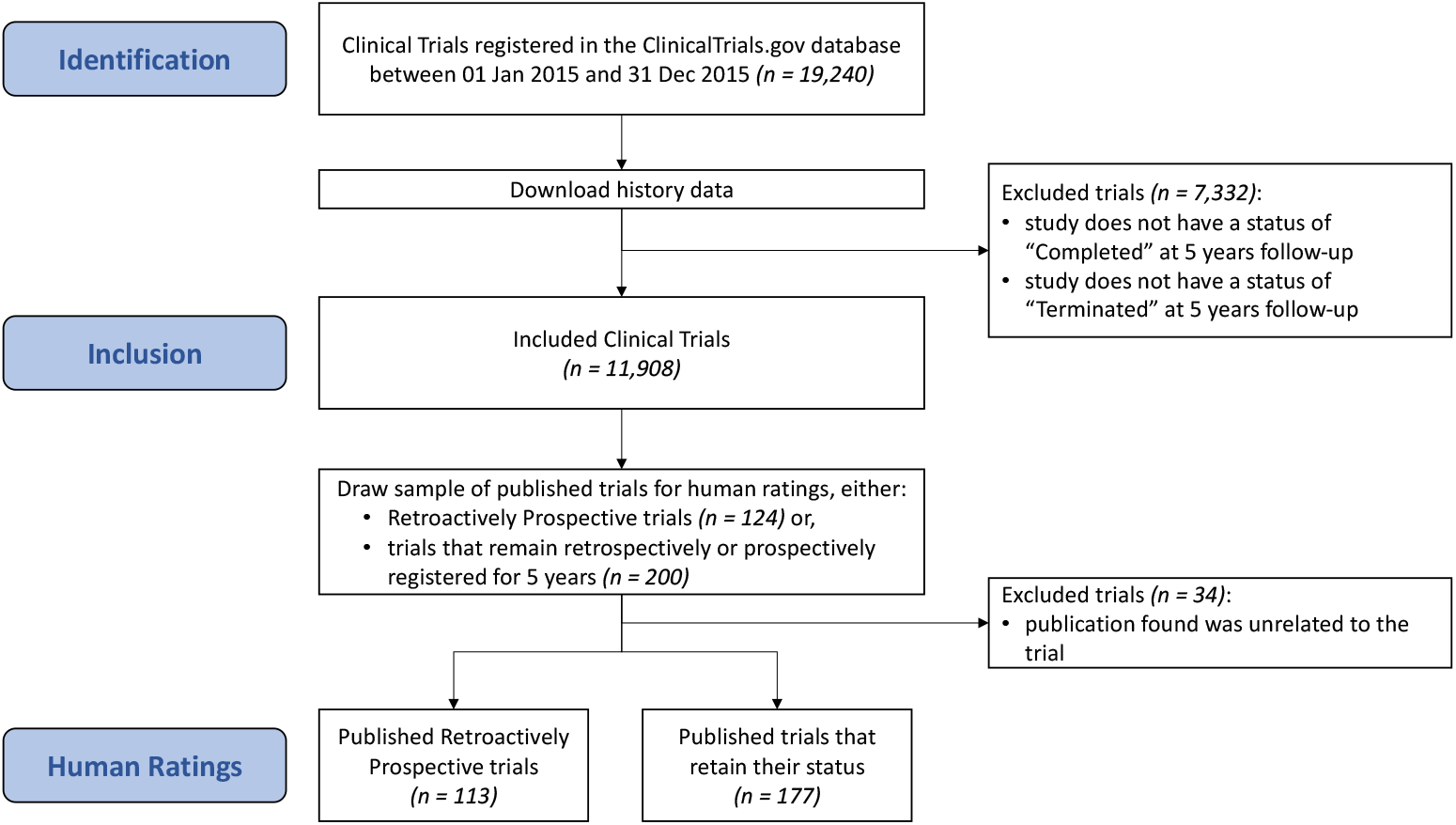
Flowchart of trials.

### Primary Objective

#### ‘Retroactively prospective’ trials

Regarding our primary objective, we found 235 trials that had been ‘retroactively prospectively’ registered, with a retrospective start date at the time of study launch, and a prospective start date at 5 years after the date of first registration. This represents 1·97% of all trials registered on ClinicalTrials.gov in 2015, thus not confirming our first hypothesis. Retroactively prospective trials account for 3·86% of all prospectively registered trials.

### Secondary Objectives

#### Post-closure ‘retroactively prospective’ trials

The majority of ‘retroactively prospective’ trials had switched their start date only after the trial had been completed (158, which is 1·33% of all trials and 67.23% of retroactively prospective trials).

#### Registration timepoints

2358 trials (19·80%) were only registered after the study end. 5843 trials (49·07%) were always registered prospectively, and 5524 trials (46·39%) were always registered retrospectively. We found a rate of 289 trials (4·97% of all retrospectively registered trials) to be ‘retroactively retrospective’, i.e., they were prospectively registered in the beginning, but changed their start date such that at 5 years, the trial was retrospectively registered.

#### Trial phases and fields

There were only few prominent differences between phases or fields. Phase 1 trials were more often prospectively registered, but also had higher rates of ‘retroactively prospective’ trials and ‘retroactively retrospective’ trials. In trials with no phase number, we found the proportion of prospective registration to be lower, while the proportions of retrospective registration and the rate of trials registered after study end were higher. In the field of cancer, there was more prospective and less retrospective registration compared to other medical fields. All further results for our primary and secondary objectives – split by field and phase number – are presented in Table 1.

**Table 1.** Numbers and proportions of trials regarding key objectives.

| Number<br>(proportion as fraction of all<br>respective trials except where<br>indicated) | all | phase |  |  | area |  |  |  |  |
| --- | --- | --- | --- | --- | --- | --- | --- | --- | --- |
|  |  | 1 | 2+ | NA | cancer | cardio | neuro | pain | other |
| Retroactively prospective clinical trials | 235<br>(1·97%) | 49<br>(2·55%) | 74<br>(1·90%) | 111<br>(1·87%) | 37<br>(2·14%) | 23<br>(1·84%) | 39<br>(2·28%) | 23<br>(2·70%) | 133<br>(1·81%) |
| Retroactively prospective clinical trials, post-closure (SO1) | 158<br>(1·33%) | 24<br>(1·25%) | 48<br>(1·23%) | 85<br>(1·43%) | 19<br>(1·10%) | 17<br>(1·36%) | 27<br>(1·58%) | 19<br>(2·23%) | 90<br>(1·23%) |
| Trials registered only after study end (SO2) | 2358<br>(19·80%) | 318<br>(16·52%) | 675<br>(17·31%) | 1334<br>(22·46%) | 206<br>(11·93%) | 232<br>(18·56%) | 273<br>(15·98%) | 182<br>(21·34%) | 1639<br>(22·31%) |
| Trials that were always registered prospectively (SO3) | 5843<br>(49·07%) | 1099<br>(57·09%) | 2171<br>(55·68%) | 2509<br>(42·25%) | 972<br>(56·28%) | 588<br>(47·04%) | 862<br>(50·47%) | 391<br>(45·84%) | 3493<br>(47·56%) |
| Trials that were always registered retrospectively (SO4) | 5524<br>(46·39%) | 714<br>(37·09%) | 1546<br>(39·65%) | 3186<br>(53·65%) | 666<br>(38·56%) | 610<br>(48·80%) | 778<br>(45·55%) | 428<br>(50·18%) | 3522<br>(47·95%) |
| Retroactively prospective clinical trials (SO5)<br>(proportion as fraction of <u>all prospective trials</u> at 5y) | 235<br>(3·86%) | 49<br>(4·26%) | 74<br>(3·29%) | 111<br>(4·24%) | 37<br>(3·67%) | 23<br>(3·76%) | 39<br>(4·33%) | 23<br>(5·56%) | 133<br>(3·66%) |
| Retroactively retrospective clinical trials (SO6)<br>(proportion as fraction of <u>all retrospective trials</u> at 5y) | 289<br>(4·97%) | 58<br>(7·50%) | 99<br>(6·00%) | 130<br>(3·92%) | 50<br>(6·97%) | 29<br>(4·54%) | 29<br>(3·59%) | 11<br>(2·51%) | 182<br>(4·91%) |

#### Concordance between original registry entries and publications

We hypothesized that among clinical trials with ‘retroactively prospective’ registration, there would be a lower rate of concordance between the originally registered study start date (post-launch) and the published start date. We tested this using a sample of 113 published ‘retroactively prospective’ and 177 control trials for which a results publication had been found and assessed by human raters, using a two-proportions z-test for equality of proportions with continuity correction. The test revealed a statistically significant difference in proportions, *χ*^*2*^(1) = 61·346, *p* = 4·787*10^−15^, with the ‘retroactively prospective’ trials having a concordance of *W* = 0·06 (6·02% of these trials mentioned the original start date in the publication), and the controls of *W* = 0·61 (60·94% of these trials mentioned the original start date in the publication). Thus, our second hypothesis was corroborated.

### Additional Analyses

#### Movements of start dates in published trials

Figure 3 shows how the start dates were moved around in our sample of published trials that were exported for human curation (113 ‘retroactively prospective’ and 177 control trials). It shows that many control trials keep their original start date, and among them, only some of the prospectively registered trials move their start dates around (which would be expected). Among the ‘retroactively prospective’ trials, however, some get moved forward by quite a bit, with the 5-year start date lying just after the first registration mark.

**Figure 3.**
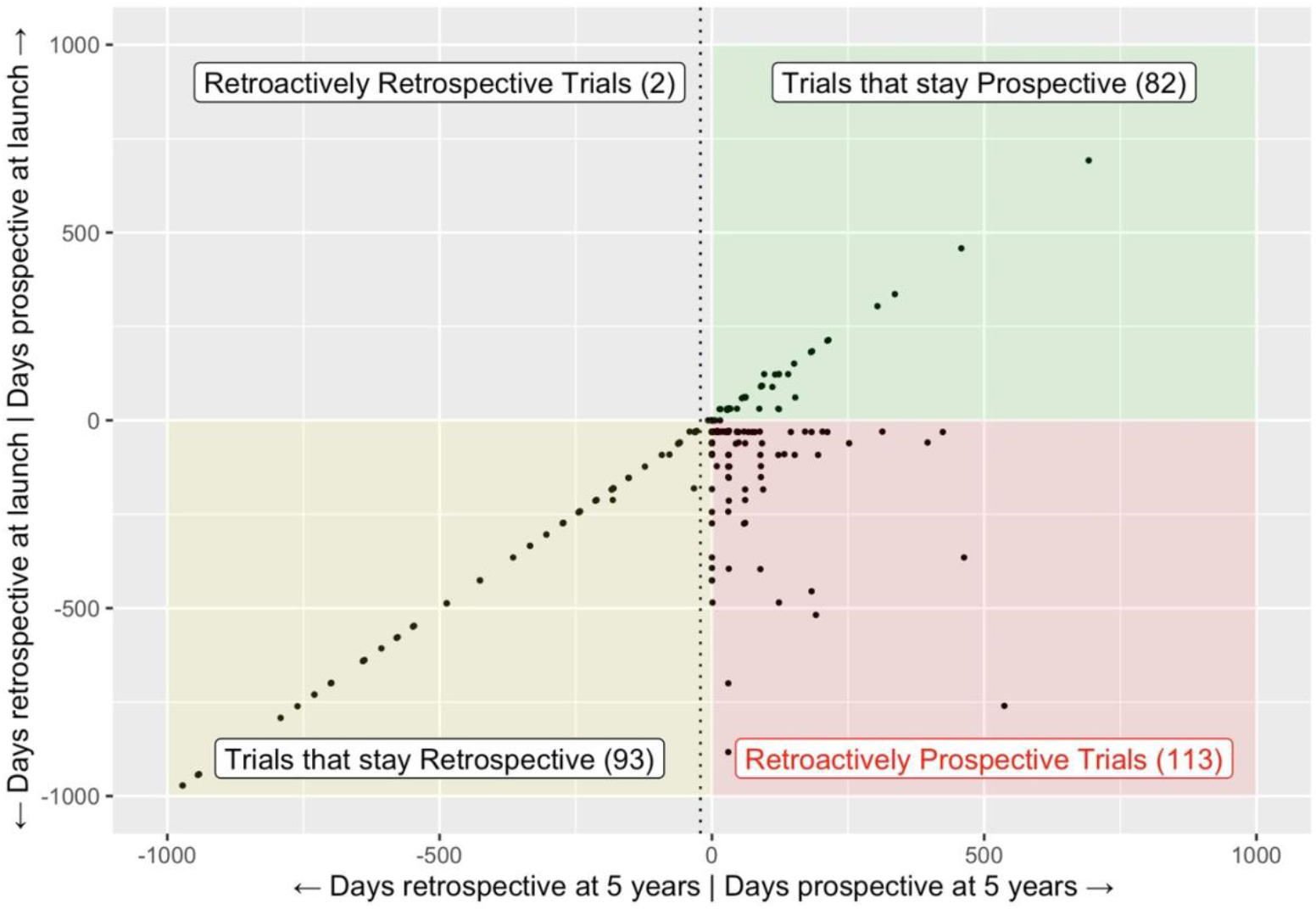
Movements of start dates from launch to 5 years, for 113 published ‘retroactively prospective’ trials and 177 published controls. The dotted line represents the 21-day ‘grace period’ granted by the FDA.

**Figure 4 (S2).**
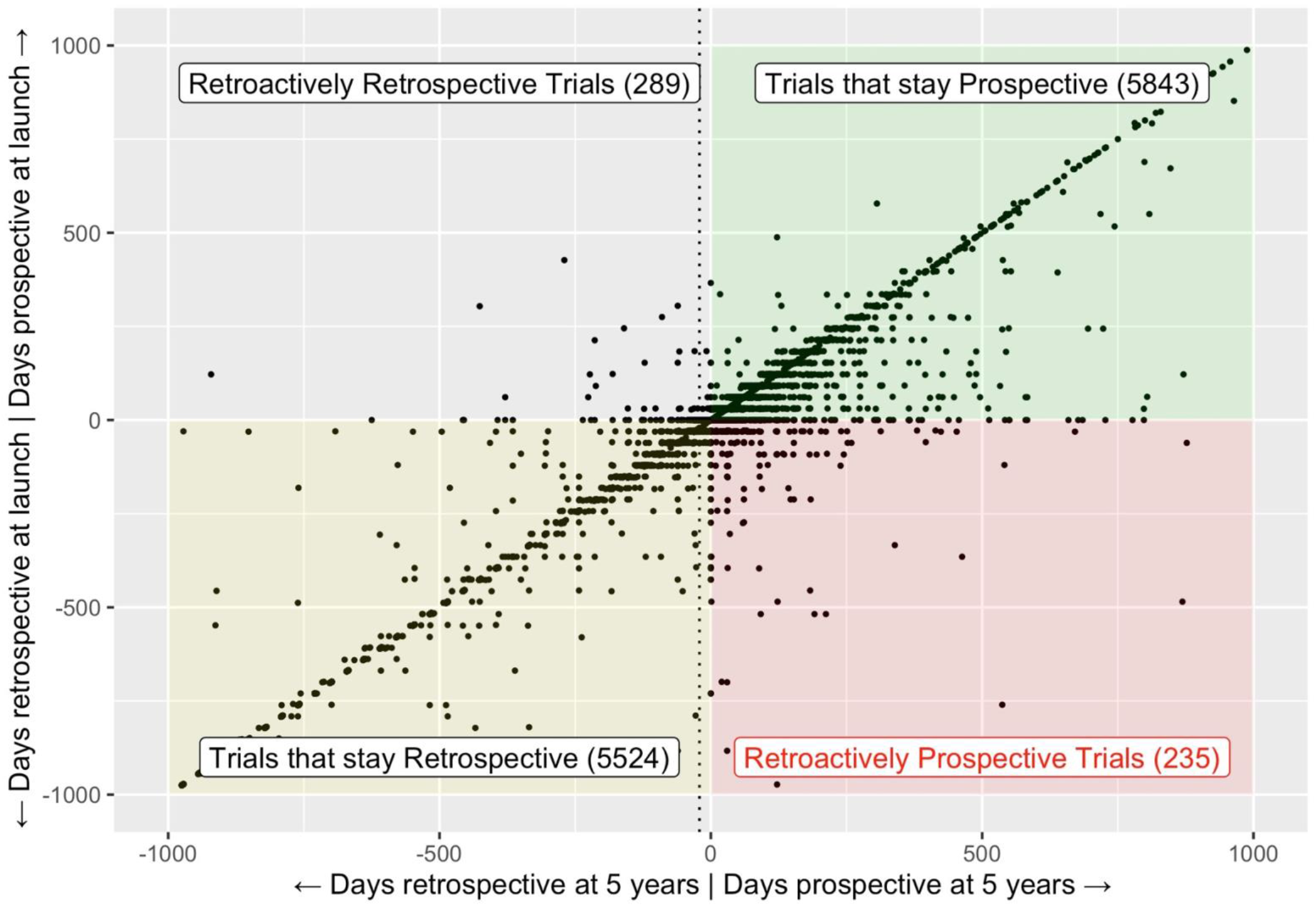
Movements of start dates from launch to 5 years, for all trials in our sample. The dotted line represents the 21-day ‘grace period’ granted by the FDA.

#### Reporting of changes in published trials

In our sample of 113 ‘retroactively prospective’ trials with an accompanying publication, none of them mentioned a change to the start date in the publication. At the same time, 12 of the ‘retroactively prospective’ trials (10·62%) explicitly stated that they were prospectively registered.

#### Movements of start dates in all trials

Analogous to Figure 3, Supplemental Figure S3 shows how the start dates were moved around in all trials in our sample. To see whether the fluctuation in the start dates is random, we calculated the median and mean of the differences: Retrospectively registered trials that changed their start date had a clear bias to the positive, with median difference of 29 days and a mean of 72·4 days (i.e., start dates got pushed forward 72 days on average).

#### Timepoints of changes to start dates in all trials

As our first secondary objective, we found that the majority of ‘retroactively prospective’ trials had switched their start date only after the trial had been completed. As additional analyses, we assessed how this compared to the proportion of all other trials with changes to their start date (i.e., trials that change their start date, but are not ‘retroactively prospective’). A two-proportions z-test for equality of proportions with continuity correction revealed a statistically significant difference in proportions, *χ*^*2*^(1) = 387·52, *p* = 2·20*10^−16^, with the ‘retroactively prospective’ trials having a proportion of *W* = 0·67 (67·23% of trials changed their start date from retrospective to prospective post-completion), and the controls having a proportion of *W* = 0·17 (17·08% of trials had their latest start date change post-completion). Additionally, we calculated at which version number the changes occur: In ‘retroactively prospective’ trials, the median version number at which the trials are changed from retrospectively registered to prospectively registered was 4, with a median of 5 versions overall.

## Discussion

Our study is the first to show that the practice of ‘retroactively prospective’ registration exists, notwithstanding the fact that our primary hypothesis was not corroborated. Almost 2 percent of trials registered in 2015 had originally been registered retrospectively and changed their start date so that by 5 years after the first registration, they appeared to have been prospectively registered. Thus, when looking at a prospective trial from 2015, there is a chance of almost four percent (3·86%) that that trial is in fact a ‘retroactively prospective’ trial. Many of these trials do not report their original start date in their publication, and none of them report a change to the start date. Twelve of these trials (10·62%) even explicitly call their study prospectively registered.

We found that Phase 1 trials had a higher rate of ‘retroactively prospective’ trials and were more often prospectively registered. This is interesting given the fact that the US Food and Drug Administration Amendments Act of 2007 Sec. 801 does not require Phase 1 trials to be prospectively registered. The practice is, however, still mandated by the ICMJE and the Declaration of Helsinki.

Our data (see Figures 2 and S2) show that the start dates fluctuate back and forth quite a bit. While it is normal for prospectively registered trials to move their start date around before the actual study start, we found that, at 5 years, some of these trials had become ‘retroactively retrospective’. We also found large fluctuations in retrospectively registered trials, which was not according to our expectations. Still, our analyses provide good evidence that fluctuations in the start dates are not just mistakes: In retrospectively registered trials, there is a clear bias toward moving the start date forward, and especially after the first-registration mark to create a ‘retroactively prospective’ trial. Also, in most cases, ‘retroactively prospective’ trials had changed their start date after study completion, and they did so within later history versions. This suggests that the changes happened to comply with editorial or regulatory policies. There are good reasons to not only register a clinical trial, but to prospectively register it. It fulfills ethical obligations, provides transparent information before the trial is launched (also to funders or regulators), and reduces publication bias. Most importantly, clinical trial registration restricts researcher degrees of freedom in the selection of hypotheses, outcomes, interventions, sampling, and makes any changes transparent. A trial that is registered prospectively would probably garner more trust than a trial that was retrospectively registered, as in the latter case, alterations to the study plan might have occurred in the period between study start and first registration, biasing the results. A trial, however, that poses as a prospectively registered trial while actually being retrospectively registered not only fails to fulfill the aforementioned obligations, but would also cover up these forms of bias. People reading the accompanying publication might get a false impression regarding the robustness of the study.

Readers, reviewers, or editors cannot determine whether a trial is a ‘retroactively prospective’ trial from examining the first page of the clinical trial record, or even using the tools provided by ClinicalTrials.gov for mass data download. While the historical data are technically publicly available and the interested reader could visit the ClinicalTrials.gov website and review all historical versions manually, this practice would hardly be feasible. This is due to a complex website design, no available API for historical data and the sheer number of versions to be reviewed (in our sample, there was an average of 6.5 versions per trial, with the maximum number of versions being 180).

Thus, even for single trials, tools are needed to help uncover these practices. The cthist R package, which was originally developed for this project and which comes with a shiny app (https://bgcarlisle.shinyapps.io/clinicaltrialshistoryscraper/), can assist journal editors, peer reviewers, other scientists, journalists, or the public in uncovering changes to the start dates. It allows these groups to judge more appropriately the bias in a trial and to make appropriate decisions or require disclosures. There is no technical reason why automated solutions that highlight this practice could not be implemented. On the one hand, clinical trial registries could implement solutions to identify and mark these trials, for example, with a badge that says ‘retrospective registration’ or ‘prospective registration’ based on the original start date. Journals, on the other hand, could implement new editorial policies that require editors, peer reviewers, or even specialized personnel to assess the registration status of submitted clinical trials. So far, reviewers check the registry entry of a clinical trial only in the minority of cases (12).

### Limitations

*3*This study is limited in that we only consider trials from ClinicalTrials.gov, which was chosen for its accessibility to historical versions, and because it is by far the largest clinical trial registry. While this allowed us to cover many clinical trials, our findings might not be generalizable to other trials in other registries. Additionally, we limited our search to trials registered in 2015, to ensure an equal follow-up of 5 years. We thus cannot provide any data for other years, or for the development over time. The practice of ‘retroactively prospective’ trial registration might have become less widespread over the years (with more and more journals following ICMJE recommendations, (13)), stayed the same, or might even have become more common. Our method for matching trials to publications depends on journal publications correctly indexing their trial number in PubMed or authors disclosing their trial number in the abstract in accordance with CONSORT (7,14). This means that trial publications that were not correctly indexed would not have been included, which might have influenced the results of our sample. Also, we did not allow any ‘grace period’, counting a trial as retrospective if it had been registered even just one day after study start. US law, for example, allows for a 21-day grace period (15). Our power calculation resulted in a sample size of 200 per group, based on expected proportions of 10% and 20%. We were, however, unable to reach this sample size, because the number of ‘retroactively prospective’ trials was lower than expected. However, our effect turned out to be much larger than expected, still allowing us to find highly significant results.

### Outlook

Future research might address the interesting finding that study start dates fluctuate by quite a bit, with trials also having their start dates pushed backwards, in some cases leading to ‘retroactively retrospective’ trials. Upcoming studies could conduct a survey, asking researchers for the reasons they had to change the start dates, or more broadly, other critical details of the registered study protocol. Studies that have dealt with changes within the registry, or between registry entries and publications (16–22), have to our knowledge not yet surveyed the authors. While we only considered the case of potential questionable research practices that concern changes to a clinical trial’s start date, similar potential issues exist, as other elements of a trial’s registration can also be changed retroactively. This includes trial outcomes, patient enrolment, eligibility requirements, and other aspects (16,17). We chose to analyze registration and start dates because it can be automated, but other registry entry changes could also be similarly analyzed.

### Conclusion

To conclude, our study is the first to shine light on the potentially questionable practice of ‘retroactively prospective’ registration. This practice undermines clinical trial transparency and integrity; however, it could be addressed with technical solutions and editorial policies. More than 10% of the retroactively prospective trials with matched publications explicitly indicated that they were prospectively registered and changes to the registered start date were not disclosed in any of these cases. While there are legitimate reasons to change a trial’s registration information, disclosure of such changes will foster confidence and encourage accuracy in clinical trial registrations.

## Data Availability

All data which are necessary to reproduce our results are available online at the Open Science Framework. Our analysis code can be found at the Codeberg repository.

https://osf.io/82vfn/

https://codeberg.org/bgcarlisle/RetroProspectiveCTR

## Acknowledgements

The authors would like to thank Daniel Strech for his support with this project and Samruddhi Yerunkar for her help with data extractions. We would also like to thank her, along with Delwen Franzen, Martin Haslberger and Nicole Hildebrand, for helpful feedback on a first draft of the manuscript.

